# Epigenome-wide association study of human frontal cortex identifies differential methylation in Lewy body pathology

**DOI:** 10.1101/2021.10.07.21264552

**Authors:** Lasse Pihlstrøm, Gemma Shireby, Hanneke Geut, Sandra Pilar Henriksen, Annemieke J.M. Rozemüller, Jon-Anders Tunold, Eilis Hannon, Paul Francis, Alan J Thomas, Seth Love, Netherlands Brain Bank, Jonathan Mill, Wilma D.J. van de Berg, Mathias Toft

**Author notes:** Corresponding author at: Department of Neurology, Oslo University Hospital, PO Box 4950 Nydalen, 0424 Oslo, Norway.

## Abstract

Parkinson’s disease (PD) and dementia with Lewy bodies (DLB) are closely related progressive disorders with no available causal therapy, neuropathologically characterized by intraneuronal aggregates of misfolded α-synuclein. To explore the role of DNA methylation changes in PD and DLB pathogenesis, we performed an epigenome-wide association study (EWAS) of 322 postmortem frontal cortex samples and replicated results in an independent set of 219 donors. We report novel differentially methylated replicating loci associated with Braak Lewy body stage near *SFMBT2, PHYHIP, BRF1*/*PACS2* and *DGKG*. The *DGKG* locus also showed evidence of DNA methylation changes in the earliest, preclinical stage of disease. Differentially methylated probes were independent of known PD genetic risk alleles. Meta-analysis provided suggestive evidence for a differentially methylated locus within the chromosomal region affected by the PD-associated 22q11.2 deletion. Our findings elucidate novel disease pathways in PD and DLB and generate hypotheses for future molecular studies of Lewy body pathology.

## Introduction

Parkinson’s disease (PD) and dementia with Lewy bodies (DLB) are progressive and debilitating neurodegenerative disorders with complex aetiology and overlapping clinical and neuropathological features. These disorders make up the second most common cause of neurodegeneration and dementia after Alzheimer’s disease (AD), and as their prevalence increases with longer life expectancy, the current estimate of 6 million PD patients worldwide is anticipated to more than double by 2040.^1,2^ Current treatment for PD and DLB is merely symptomatic, and improved understanding of the molecular disease mechanisms is urgently needed in order to facilitate the development of targeted disease-modifying therapy. Over the last decade, large-scale genome-wide association studies (GWAS) have identified an increasing number of common genetic risk loci for PD^3^, and more recently also for DLB^4,5^, providing novel insights into pathogenesis. In contrast, previous studies have only explored to a limited degree how disease susceptibility and progression may be shaped by epigenetics. Epigenetic processes contribute to variability in complex traits such as aging and disease^6,7^ and involve mechanisms of gene regulation that are partially dynamic over time, cell-type specific and influenced by both genetic and environmental factors.

DNA methylation at CpG dinucleotides is the most frequently studied epigenetic mechanism in complex disease. Previous EWAS analyses of AD-associated neuropathology have identified replicable and consistent patterns of differentially methylated CpGs across the genome in postmortem human brain tissue, contributing important insights to disease mechanisms.^8-14^ In contrast, similar efforts in PD or DLB have thus far only been reported for very limited sample sizes^15-18^, although larger EWAS in PD have been performed on whole blood.^19-21^ A recent study investigated frontal cortex neurons isolated by a flow cytometry-based approach and detected widespread differences in DNA methylation between 57 PD patients and 48 controls, with replication in a smaller dataset, highlighting dysregulation of *TET2* in particular.^22^

Lewy bodies and Lewy neurites are intraneuronal aggregates composed mainly of misfoldedα-synuclein protein and constitute the neuropathological hallmark lesions of both PD and DLB.^23^ According to the staging system proposed by Braak^24^, Lewy pathology in PD spreads in a predictable pattern over the course of the disease from the olfactory bulb and brainstem (Braak Lewy body stages 1-3) in a caudal-to-rostral manner, next to limbic regions (Braak Lewy body stage 4), reaching the neocortex in the last stages (Braak Lewy body stages 5 and 6). Only the retrospective information about clinical symptom progression distinguishes late-stage PD from DLB, where cortical Lewy pathology and associated cognitive symptoms are already present early in the disease course.^25^ Of note, cellular and proteomic studies have demonstrated that neuronal stress and dysfunction are evident in the cortex also at the very early stages of PD-related pathology, long before Lewy bodies appear in the same brain region.^26^ Previous transcriptome studies on post-mortem human brain tissue across Braak Lewy body stages have also highlighted early changes, involving deregulation of the immune response, axonal signalling and endocytosis.^27,28^ We hypothesized that epigenetic dysregulation contributes to the common molecular pathogenesis of Lewy body disorders, and that an increasing degree of disease-related changes should be detectable in frontal cortex as the disease progresses into more advanced stages.

To dissect the role of DNA methylation changes in PD and DLB, we performed a methylome-wide association study of 322 postmortem frontal cortex samples with Braak Lewy body stages 0-6 and replicated our findings in an independent dataset from 219 donors. The size and direction of effects for the most associated probes were highly correlated across the two datasets, providing a strong indication of consistent results. We report 4 replicating CpGs significantly associated with Braak stage, explore these loci in relation to known genetic risk factors and publicly available data and provide a benchmark for further epigenetic studies of post-mortem human brain tissue in Lewy body disorders.

## Results

### Identification of differentially methylated CpGs across Braak α-synuclein stages

For the discovery stage of our analysis, we used superior frontal gyrus gray matter tissue samples from the Netherlands Brain Bank (NBB) and Normal Aging Brain Collection, Amsterdam (NABCA), including neurologically healthy controls (n = 73), donors without clinical neurological symptoms but with incidental Lewy body disease at autopsy (iLBD) (n = 29), neuropathologically confirmed PD patients (n = 139) and DLB patients (n = 81) (Supplementary Table 1, Supplementary Figure 1). Samples were screened for monogenic causes of parkinsonism using the Illumina NeuroChip and no definitely or probably pathogenic variants were discovered in *SNCA, LRRK2, VPS35* or other relevant neurodegenerative genes covered by the array.^29^ Genome-wide DNA methylation was assessed using the Illumina Infinium MethylationEPIC array. Following stringent quality control and filtering (see Online Methods), data on 583,192 CpG probes in 322 individuals were included in statistical analyses. We used a published algorithm and reference data to estimate the proportion of NeuN positive neurons in each tissue sample and included this variable as a covariate, as well as sex, age, postmortem interval, experiment plate and the first three surrogate variables (see Online Methods). We observed a non-significant trend towards lower estimated proportion of neurons in samples with higher Braak Lewy body stage (Pearson r^2^ -0.065, p = 0.24).

In the discovery stage, 24 CpG probes were associated with Braak α-synuclein stage at FDR < 0.05 (Supplementary Table 2). Next, we used equivalent inclusion criteria and analysis pipeline to select samples for a replication dataset from 219 donors in the UK Brains for Dementia (BDR) cohort with DNA methylation data available.^30^ Of note, the distribution of individuals across Braak Lewy body stages 0-6 was different across the two datasets, with a higher proportion of neuropathologically healthy donors in the replication cohort (68% vs 23% Braak stage 0) (Supplementary table 1). We observed that DNA methylation differences across Braak Lewy body stages were strongly correlated across the two datasets for the 24 FDR-significant sites identified in the discovery phase (Pearson r^2^ 0.65, p = 0.00063, 79% concordant direction, binomial sign test p = 0.0066) (Figure 1). Associations for 4 DNA methylation sites were significantly replicated in the BDR dataset (two-sided p < 0.05 and same direction of effect) including cg14511218 (not annotated to any coding gene, located downstream of *SFMBT2*), cg04011470 (annotated to *PHYHIP*), cg19898425 (annotated to *BRF1*/*PACS2*) and cg13986157 (annotated to *DGKG*) (Table 1, Figure 2).

**Figure 1.**
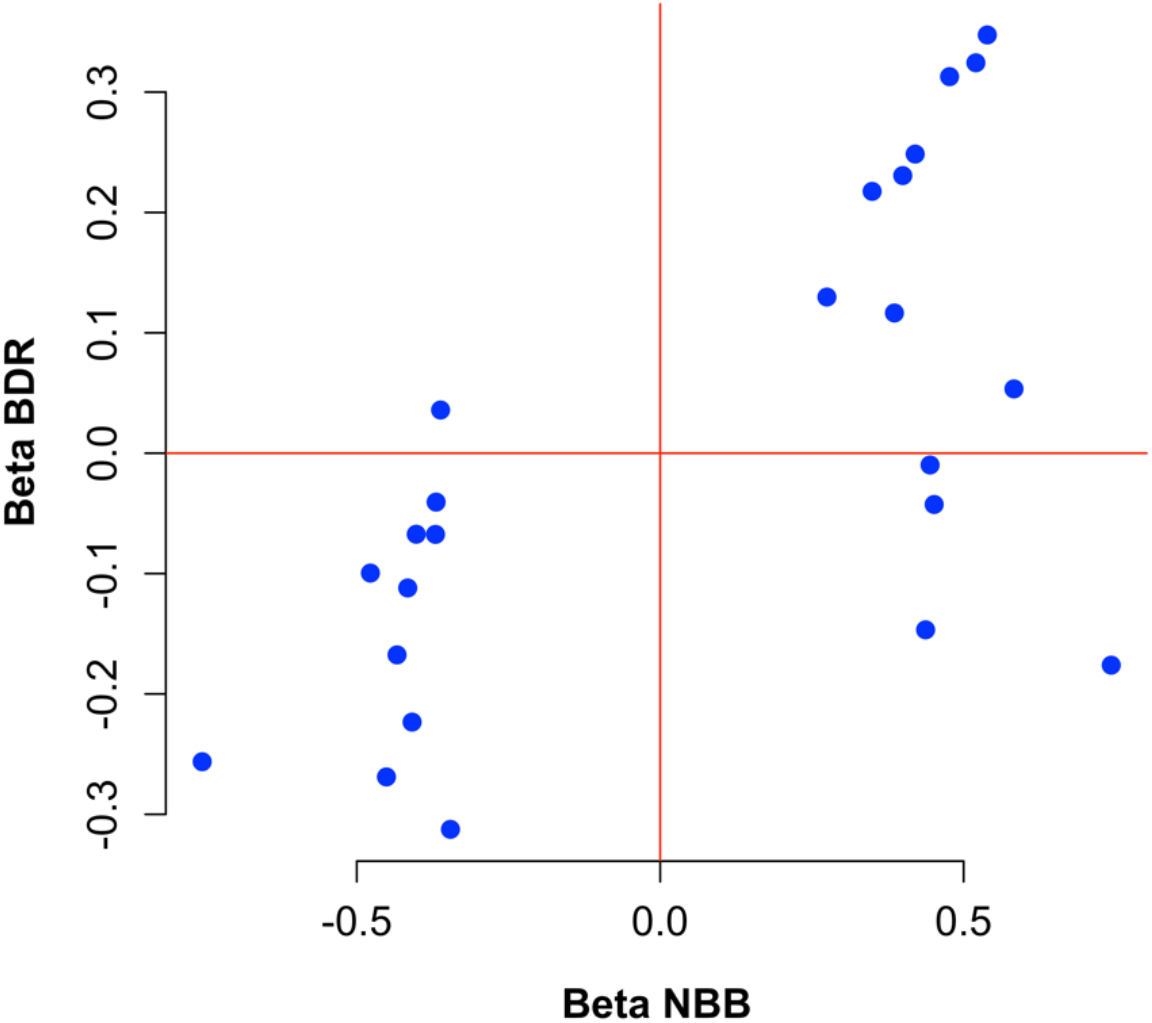
Correlation between effect size in discovery and replication data for top probes. The figure shows the beta values from linear regression for the 24 probes passing a false discovery rate threshold in the discovery stage. A strong correlation between the discovery (NBB, Netherlands Brain Bank) and replication (BDR, Brains for Dementia Research) analyses indicates consistent results.

**Table 1.**
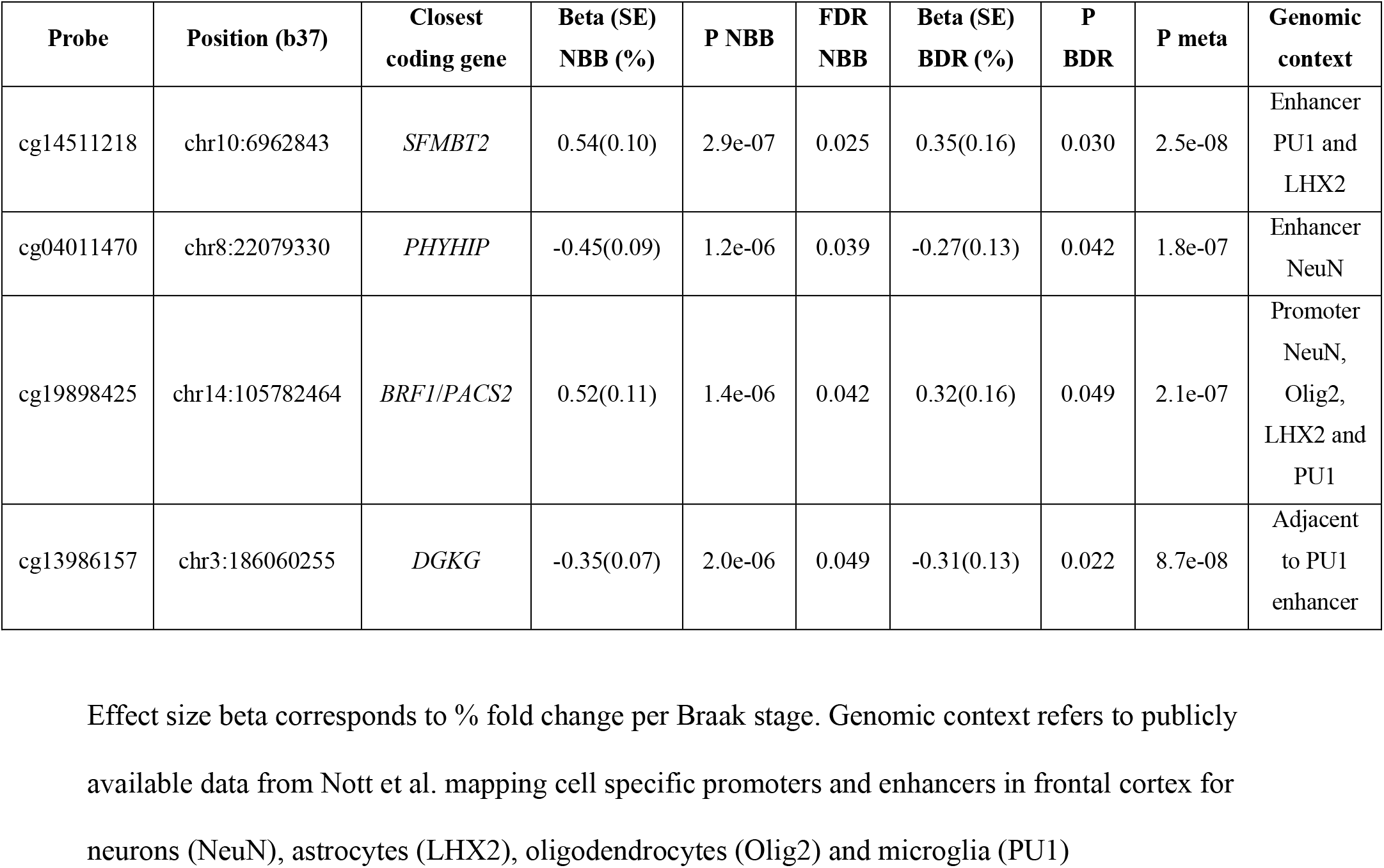
Replicating CpG probes associated with Braak Lewy body stage.

| Probe | Position (b37) | Closest coding gene | Beta (SE) NBB (%) | P NBB | FDR NBB | Beta (SE) BDR (%) | P BDR | P meta | Genomic context |
| --- | --- | --- | --- | --- | --- | --- | --- | --- | --- |
| cg14511218 | chr10:6962843 | *SFMBT2* | 0.54(0.10) | 2.9e-07 | 0.025 | 0.35(0.16) | 0.030 | 2.5e-08 | Enhancer<br>PU1 and<br>LHX2 |
| cg04011470 | chr8:22079330 | *PHYHIP* | -0.45(0.09) | 1.2e-06 | 0.039 | -0.27(0.13) | 0.042 | 1.8e-07 | Enhancer<br>NeuN |
| cg19898425 | chr14:105782464 | *BRF1/PACS2* | 0.52(0.11) | 1.4e-06 | 0.042 | 0.32(0.16) | 0.049 | 2.1e-07 | Promoter<br>NeuN,<br>Olig2,<br>LHX2 and<br>PU1 |
| cg13986157 | chr3:186060255 | *DGKG* | -0.35(0.07) | 2.0e-06 | 0.049 | -0.31(0.13) | 0.022 | 8.7e-08 | Adjacent<br>to PU1<br>enhancer |

Effect size beta corresponds to % fold change per Braak stage. Genomic context refers to publicly available data from Nott et al. mapping cell specific promoters and enhancers in frontal cortex for neurons (NeuN), astrocytes (LHX2), oligodendrocytes (Olig2) and microglia (PU1)

**Figure 2.**
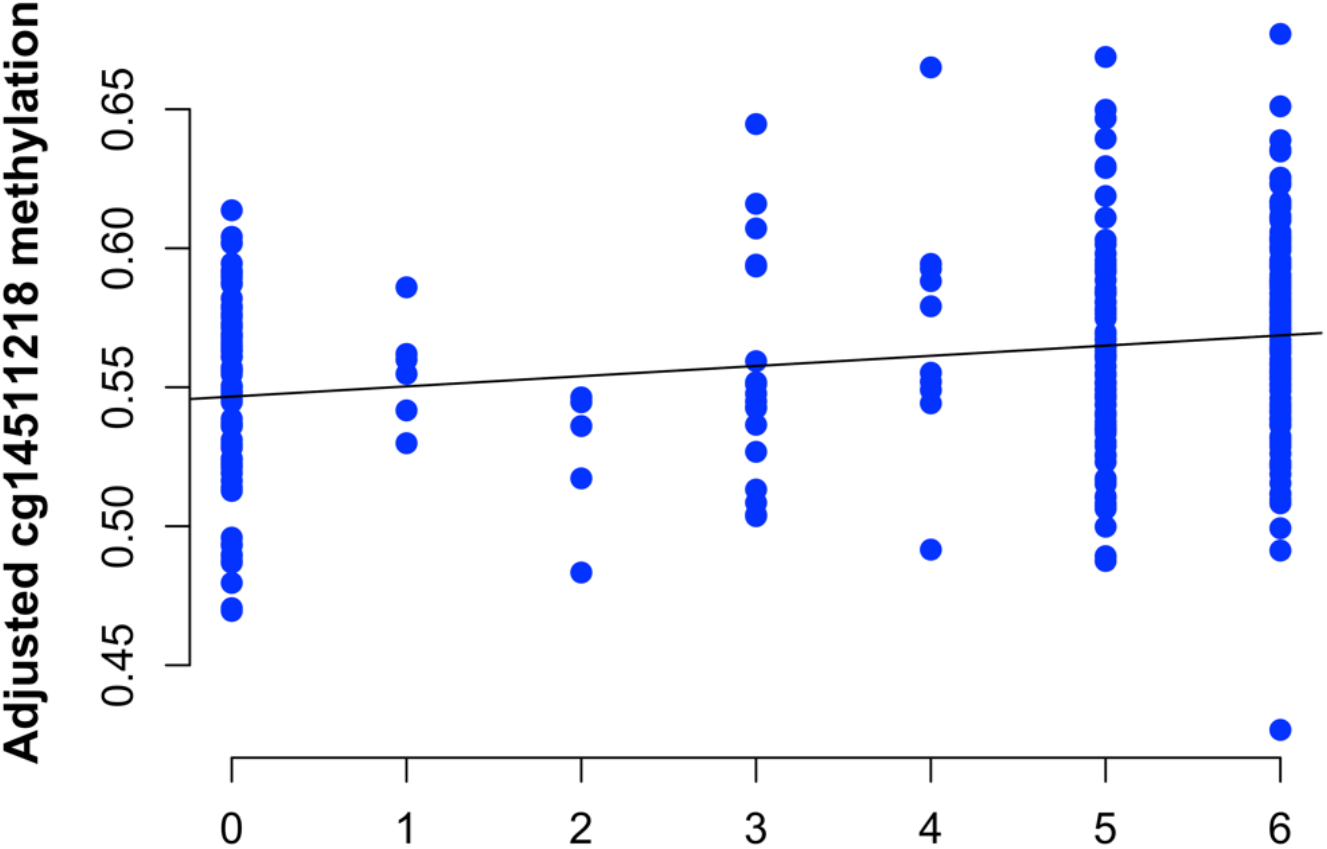
Adjusted methylation values for the significant probe cg14511218 across Braak stages. The figure shows individual methylation values in the discovery dataset, adjusted for covariates as described in the methods section. A regression line is shown indicating increasing methylation levels with higher Braak α-synuclein stage.

In addition to the two-stage discovery and replication approach, we also meta-analyzed the 566,306 probes passing QC in both datasets. In a fixed-effect inverse variance meta-analysis, 54 probes were significant at FDR < 0.05 and 28 of these were associated at p < 0.05 in both datasets with the same direction of effect (Supplementary Table 3). As expected, the significant DNA methylation sites identified by the two-stage approach were also high on the list of 28 probes highlighted via meta-analysis, representing 4 out of the 5 lowest p-values. In addition, cg03318382 (not annotated to any coding gene, located upstream of *SEPTIN5*) emerged with the second strongest p-value in the dataset. This probe was not FDR-significant in the discovery stage alone (NBB p = 1.8*10^−5^, beta = 0.46), but a larger relative effect size in the replication dataset (BDR p = 0.0011, beta = 0.52) resulted in a strong association in the meta-analysis (meta-analysis p = 5.0*10^−8^, beta = 0.48). In meta-analysis, the three most significant probes also passed the more conservative Bonferroni-corrected threshold (p < 8.8*10^− 8^, correcting for 566,306 tests).

### Exploring the genomic context of differentially methylated CpGs

Sites profiled by the MethylationEPIC array primarily map to promoters and enhancers in the human genome where variability in CpG methylation is likely to affect the functional regulation of nearby genes. To potentially nominate genes and cell-types implicated by our Braak Lewy body stage associated CpGs, we assessed the overlap between probe positions and genomic annotations from a recent publication characterizing the noncoding regulatory regions and enhancer-promoter interactome of major cell types in the human cortex.^31^ In this previous work, active promoters and enhancers were identified by assay for transposase-accessible chromatin sequencing (ATAC-seq) and chromatin immunoprecipitation sequencing (ChIP-seq) for histone modifications H3K27ac and H3K4me3. Table 1 includes a summary of probe position overlap with promoters and enhancers in neurons, microglia, astrocytes and oligodendrocytes.

The strongest associated replicating probe, cg14511218, is located ∼240kb downstream of *SFMBT2* on chromosome 8, where there is evidence of enhancer activity in microglia and astrocytes, but most pronounced in microglia. Data from proximity ligation-assisted chromatin immunoprecipitation sequencing (PLAC-seq) in cortical microglia indicate that the enhancer makes its strongest interactions with the *SFMBT2* promoter.^31^ Further evidence of the relevance of microglia to the cg14511218 association comes from the inclusion of *SFMBT2* in the human brain “microglial signature” published by Gosselin *et al*., comprising 881 transcripts showing minimum 10-fold increased expression in microglia relative to cortex tissue.^32^ *SFMBT2* is also among the microglial genes highlighted as being differentially expressed in PD relative to control brains in a previous study.^33^

Cg04011470 is located in the last of 4 coding exons of *PHYHIP*. Epigenetic marks indicate a neuronal enhancer at the locus, interacting with several other genes in the region, some of which are also considered candidate genes for the *BIN3* PD GWAS association signal. The third differentially methylated CpG, cg19898425, is in the first intron of *PACS2*, corresponding to the promoter region in all major brain cell types. Cg13986157 maps near a microglial enhancer in intron 1 of *DGKG* (Table 1).

The DNA methylation site nominated primarily from meta-analysis, cg03318382, is located ∼13kb upstream of *SEPTIN5*, not overlapping with a promoter or enhancer for any brain cell type. We note, however, that its location on chr22:19691974 (GRCh37) falls within the ∼3Mb region typically affected in the 22q11.2 deletion syndrome, which has been implicated in a number of clinical phenotypes, including idiopathic PD.^34^

### Association between CpG methylation and preclinical Lewy body pathology

End-stage neurodegeneration implies profound structural and functional disturbances. In samples from donors with advanced disease, changes that play a relevant causal role in the pathogenic process are not easily distinguished from general downstream consequences, posing a challenge for the interpretation of postmortem experiments. To assess potential early changes, we analysed DNA methylation levels for the 4 significant CpG probes in a subset of discovery samples, contrasting neurologically healthy donors with iLBD (N = 29) and without iLBD (N = 73). Lewy body pathology is found in approximately 10% of healthy routine postmortem examinations, and evidence of accompanying neuronal loss supports the notion that iLBD represents presymptomatic PD.^35^ Despite the limited sample size, one of the probes (cg13986157) showed significant differential methylation comparing iLBD and non-LBD controls (two-sided p = 0.039), with DNA methylation in the iLBD group at an intermediate level between neurologically healthy controls and clinically manifest Lewy body disease (Figure 3).

**Figure 3.**
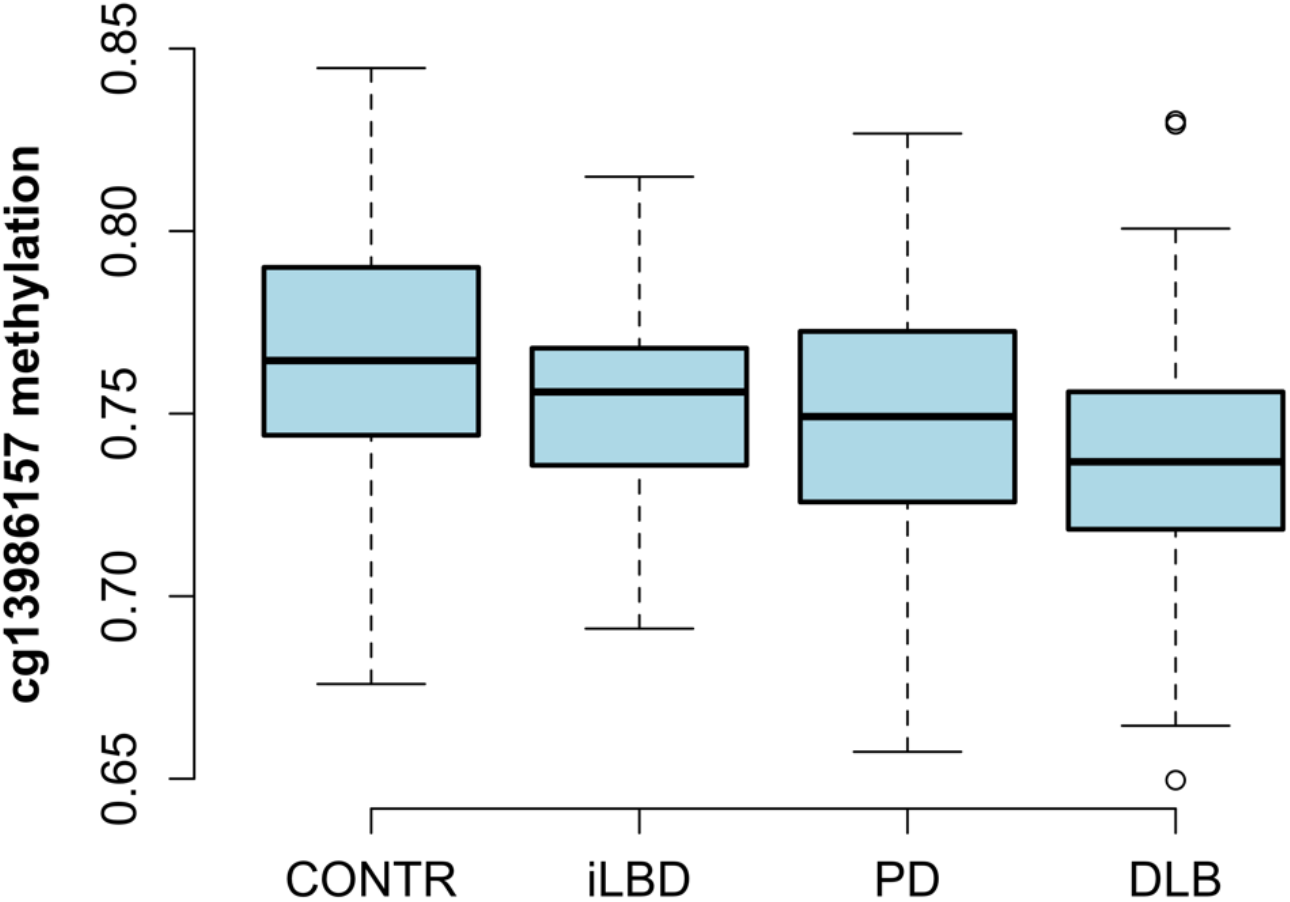
Boxplot of cg13986157 methylation across neuropathological diagnoses. The plot shows methylation levels across diagnostic groups for cs13986157. Error bars represent 1.5 times the interquartile range. The difference betweeen controls and iLBD is significant at two-sided p = 0.039.

**Figure 4.**
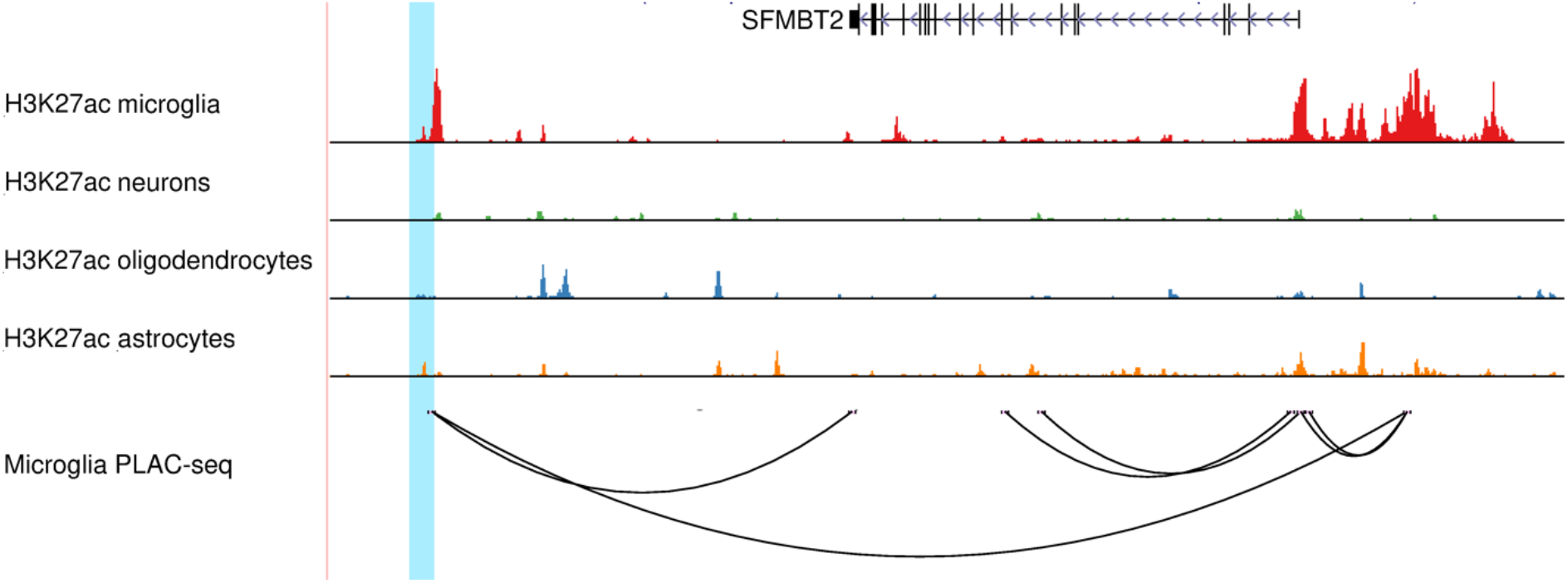
The brain-specific genomic context of cg14511218 and relation to *SFMBT2*. The figure shows chromosome 10, position 6,000,000 - 7,600,000 (b37), with the location of cg14511218 highlighted. Data from Nott et al. show histone acetylation associated with active enhancers and promoters (H3K27ac) across different cell types, and proximity ligation-assisted chromatin immunoprecipitation sequencing (PLAC-seq) demonstrating enhancer-promoter contact in microglia.

### Analyses of known genetic risk loci and methylation-quantitative locus analysis

Of note, one of the significant CpG probes (cg04011470) is located less than 500kb from rs2280104 near *BIN3*, a genome-wide significant PD risk locus.^3^ Comparing DNA methylation and genotype data in our discovery sample, we found no association between cg04011470 and rs2280104, nor with any other single-nucleotide polymorphism (SNP) within a 1Mb window around the probe position. Similarly, we found no significant methylation-quantitative loci (mQTL) for cg04011470 among 277 cis-SNPs in publicly available frontal cortex data summary statistics from 543 individuals in the Brain xQTL server.^36^

Turning to other known risk loci for PD, we extracted results from all 26,743 CpG probes located within 500kb of any significant GWAS SNP reported by Nalls et al., but cg04011470 remained the only association significant when correcting for multiple testing. Hypomethylation of the *SNCA* promoter region in PD has been reported in a number of previous studies, both in brain and blood.^37-39^ We therefore assessed CpG probes in the *SNCA* locus specifically, but found no significant CpGs when correcting for multiple testing of 109 probes within 500kb of significant SNPs (minimum p-value 0.0035 for cg01035160).

Next, we assessed genotype data for cis-mQTLs within 500kb of our remaining 3 Braak stage-associated CpGs, but found no SNPs significantly associated with methylation levels for any of the probes. These results suggest that the DNA methylation differences associated with Lewy body pathology do not appear to be mediated by underlying genetic variability.

## Discussion

We performed a methylome-wide association study across Braak Lewy body stages in postmortem human frontal cortex, and followed up our findings in an independent dataset, identifying 4 novel differentially methylated loci that replicated across studies. Including a total of 541 participants, we analysed a larger sample than the few previously published EWAS studies of human brain tissue in Lewy body disorders.^17,18^ Our study design mirrors a series of successful bulk cortex EWAS studies of AD neuropathology, where the number of significant CpG associations has increased from a handful in the first published studies^8,9^ to several hundred in recent meta-analyses^10,14^, providing novel insights into disease pathways and mechanisms. We demonstrate here for the first time that the same approach is feasible also for Lewy body pathology, the hallmark of PD and DLB.

Differentially methylated sites are located in promoters or enhancers, and interpretations linking association signals to specific genes should be made with caution in the absence of functional evidence. Based on genomic annotations from human cortex, we raise the hypothesis that dysregulation of *SFMBT2* in microglia may be linked to disease. *SFMBT2* encodes Scm-like with four malignant brain tumor domains 2. Apart from being reported as a microglial signature gene^32^ and differentially expressed in PD^33^, its function in the human brain is largely unexplored.

Hypermethylation of cg19898425 within the *PACS2* promoter also nominates this gene as potentially implicated in Lewy body disorders. The corresponding protein, phosphofurin acidic cluster sorting protein 2, regulates interactions at mitochondria-associated endoplasmatic reticulum membranes, a mechanism that can be linked to a number of key pathways and molecular players in PD pathogenesis, such as calcium signalling, mitochondrial dynamics, inflammation, and autophagy, making *PACS2* an interesting candidate gene for further study.^40^

Differentiating causes from effects is a constant challenge in epigenetic studies of complex disease, and particularly difficult for brain disorders, where longitudinal sampling is impossible for the main tissue of interest.^41^ However, taking advantage of samples from donors with iLBD gave us some opportunity to evaluate potential preclinical changes in a case-control design. Despite the limited sample size, we found that cg13986157 in *DGKG* was significantly hypomethylated in iLBD cases compared to neurologically healthy controls without iLBD, indicating that at least some disease-associated methylation changes are likely to occur early in the development of Lewy pathology, even before symptoms manifest.

We noted with interest that the differentially methylated probe cg04011470 in *PHYHIP* was located less than 500kb from the top associated SNP identified in a recent PD GWAS analysis. Studies of the association between common SNPs, gene expression, histone modifications and CpG methylation in the human brain have revealed that the different types of QTLs show a certain degree of overlap and are all enriched among disease-associated GWAS SNPs^36^, raising the possibility that our finding could be linked to the same mechanism as the adjacent GWAS signal. However, in the same study, SNP effects on RNA expression were only mediated fully by epigenetic variation in 9% of loci, so simple one-to-one relationships remain the exception. Consequently, we are not surprised that we did not identify significant mQTLs for cg04011470 or any of the other differentially methylated CpGs, which is also in line with a recent large meta-analysis of AD neuropathology EWAS.^10^ The lack of mQTLs is still compatible with a hypothesis that genetic and epigenetic association signals from the same genomic region may represent independent disease mechanisms converging on the same gene.

In meta-analysis, we identified 28 CpGs associated with Braak Lewy body stage with consistent direction of effect and p < 0.05 in both the NBB and the BDR datasets. This approach is less conservative than the standard two-stage discovery and replication method, and should therefore be interpreted with caution. The strongest additional signal emerging in our meta-analysis was an association between Lewy pathology and hypermethylation of cg03318382, located within the ∼3Mb region typically affected in the 22q11.2 deletion syndrome. This deletion occurs in at least 1 in 4000 births and has been linked to a number of clinical phenotypes including DiGeorge syndrome, velocardiofacial syndrome and psychotic illness.^42^ Early-onset parkinsonism reported as a clinical feature in known carriers of the 22q11.2 deletion sydrome^43^ prompted further investigations confirming its role as a risk factor for PD, also predisposing to early onset.^34,44^ It is not clear which genes within the region are important for PD pathogenesis, although *COMT*, encoding cathecol-o-methyltransferase has been highlighted as an interesting candidate.^34^ We found CpG hypermethylation upstream of *SEPTIN5*, which encodes septin 5. Septins are guanosine-5’-triphosphate (GTP)-binding proteins that contribute to the regulation of synaptic vesicle trafficking and neurotransmitter release, and septin 5 has been shown to interact with parkin, a protein implicated in early-onset autosomal recessive PD.^45^ Our findings highlight the possibility that epigenetic changes in the 22q11.2 region may be associated with disease risk in individuals with normal copy number due to dysregulation of one or more of the same genes that contribute to the 22q11.2 deletion sydrome.

Hypomethylation at the promoter in intron 1 of *SNCA* has been reported in postmorten human brain. One study showed significant differences in both substantia nigra and cortex^37^, whereas another found significant change in substantia nigra only^38^, with similar methylation levels in patients and controls in putamen and the anterior cingulate. Furthermore, *SNCA* hypomethylation has been demonstrated in whole blood and cultured mononuclear cells, also with an mQTL association to susceptibility SNPs for PD and DLB in brain and blood^39,46^, but the methodologies used have been different from the present study. A number of factors, including choice of brain region, array design and adjustment for cell composition may have contributed to our inability to replicate this signal.

Our study has several limitations. The standard sodium bisulfite conversion method used in MethylationEPIC array analysis does not capture variability in hydroxymethylation, an intermediary DNA modification state between methylated and unmethylated cytosine that is enriched in the brain and serve specific regulatory functions.^47^ Parallel profiling of methylation and hydroxymethylation has been shown to improve the resolution and interpretation of epigenetic studies in AD.^48^ Depending on the availability of tissue and resources, we investigated only superior frontal cortex, where we ideally would have assessed tissue from multiple brain regions. Experience from AD EWAS indicate some regional variation in differential DNA methylation, yet substantial overlap between significant CpGs across different parts of the cerebral cortex.^10^ Substantia nigra is particularly vulnerable to Lewy pathology and pivotal in PD pathogenesis, yet postmortem methylation studies of this region is of limited value due to the profound neuronal loss. We used a published algorithm to estimate cell composition from methylation profiles and adjust for this variable in our analysis. The reference dataset only distinguishes NeuN positive neurons from non-neuronal cells, however, and even with good bioinformatic corrections, epigenetic studies of bulk tissue will never obtain a resolution comparable to investigations of pure cell populations.

A recent study isolated frontal cortex neurons by a flow cytometry-based approach and reported widespread methylation differences between 57 PD patients and 48 controls, with replication in a smaller dataset.^22^ In particular, the authors highlighted dysregulation of *TET2*, involving a pattern of promoter hypomethylation and enhancer hypermethylation. This study assessed methylation by bisulfite padlock probe sequencing, so the results are not directly comparable to our MethylationEPIC array data. We did not detect significant association signals from the *TET2* locus, but we note that only a subset of CpGs were differentially methylated in the original study, and our data did not precisely cover the same positions. We acknowledge that methylation studies in sorted cell populations have many advantages over our design, as less noisy data is both more powerful and easier to interpret. However, as bulk tissue approaches like ours remain far more scalable to large sample sizes, we anticipate that both cell type specific and bulk brain tissue EWAS designs will have important roles to play in mapping the epigenetics of Lewy body neuropathology. We also note that there is ample evidence to support a role also for non-neuronal cells in the pathogenesis of Lewy body disorders.^49,50^ The large-scale bulk tissue design has the ability to potentially pick up strong signals driven by any cell type, as our exploration of overlap between differentially methylated CpGs and functional genomic annotation for major brain cell types also indicates.

Despite being larger than previous brain EWAS in PD and DLB, the sample size of our study was still modest, limiting our statistical power for hypothesis-free association analysis. As the total number of samples have passed one thousand for meta-analyses of EWAS in AD neuropathology, a larger set of robustly associated CpGs can be utilized for enrichment analyses, linking the results collectively to molecular mechanisms and pathways. We hope to see a similar development in Lewy neuropathology EWAS through collaborative efforts in the years to come. Furthermore, cross-trait analyses are warranted to assess the degree of epigenetic overlap versus disease-specific methylation patterns across neurodegenerative disorders. Understanding causality and the temporal sequence of disease-linked processes is also of major importance and could potentially be addressed through studies in model systems as well as larger collections of postmortem samples in the very early stages of neuropathological change. Finally, more studies are needed to identify the genetic and environmental determinants driving disease-associated methylation changes in neurodegenerative disorders.

In summary, we performed the first well-powered two-stage EWAS of Lewy body neuropathology in postmortem human frontal cortex, providing evidence for significant methylation differences associated with Braak α-synuclein stages. We link novel genomic loci to the pathology underlying PD and DLB, including one CpG showing evidence of methylation change already in the preclinical phase. These findings generate hypotheses for further molecular studies and hold promise that future meta-analyses in larger sample sets will be as fruitful in Lewy neuropathology as in recent studies of AD-related changes. Complementing other genomic approaches such as GWAS and gene expression studies, investigations of DNA methylation may provide important contributions to our basic understanding of disease mechanisms and ultimately facilitate the development of disease-modifying therapy for PD and DLB.

## Online methods

### Subjects and samples

Samples in the discovery data set were obtained from the Netherlands Brain Bank (NBB, www.brainbank.nl) and Normal Aging Brain Collection, Amsterdam (NABCA).^51^ Written informed consent for the use of tissue samples and clinical information for research purposes was collected from the donors or their next of kin. The Medical Ethics Committee of the VU University Medical Centre, Amsterdam, approved all procedures of NBB and NABCA, and the study was approved by the Regional Committee for Health and Medical Research Ethics, Norway. We included samples from controls without any records of neurological or psychiatric disease during life, neurologically healthy donors with iLBD, clinically diagnosed and pathologically-confirmed PD patients and DLB patients. Deliberately aiming to focus our analysis on this spectrum of Lewy body disease, we excluded samples of donors with pathologically confirmed AD either alone or in addition to one of these diagnoses. Standardized brain autopsies and neuropathological examinations were performed by experienced neuropathologist or neuroanatomist (AR and WB), including assessment of Lewy body-related α-synuclein pathology according to BrainNet Europe guidelines.^52^ Clinical information was extracted from medical records provided by the NBB. PD diagnosis was based on the combination of clinical parkinsonism according to UK Parkinson’s Disease Society Brain Bank^53^ or Movement Disorders Society^54^ criteria and moderate to severe loss of neuromelanin-containing neurons in the substantia nigra in association with Lewy pathology in at least the brainstem, with or without limbic and cortical brain regions.^55^ Criteria for DLB were a clinical diagnosis of probable DLB according to the consensus criteria of the DLB Consortium^25^ combined with presence of limbic-transitional or diffuse-neocortical Lewy pathology upon autopsy. Dementia was diagnosed clinically during life by a neurologist or geriatrician, or retrospectively based on neuropsychological test results showing disturbances in at least two core cognitive domains^56^ or Mini-Mental State Examination (MMSE) score <20. The distinction between DLB and PD with dementia (PDD) (74 out of 139 PD patients) was made based on the “1-year rule”, classifying dementia presenting before or within 1 year of parkinsonism onset as DLB.^25^ Tissue slices of 50-100mg macroscopically spanning all cortical layers, grey matter only, were cut from frozen frontal cortex tissue blocks, collected at autopsy and stored at -80oC until further processing, in a cryostat, and DNA was extracted using the Qiagen AllPrep DNA/RNA/miRNA Universal Kit according to the manufacturer’s instructions.

A subset of donors from the BDR DNA methylation dataset^30^ were used to replicate specific DMPs from the discovery cohort. We used prefrontal cortex Illumina Infinium MethylationEPIC BeadChip data from 219 individuals. Briefly, the BDR cohort was established in 2008 and consists of a network of six dementia research centers across England and Wales (based at Bristol, Cardiff, King’s College London, Manchester, Oxford and Newcastle Universities) and five brain banks (the Cardiff brain donations were banked in London). BDR is approved as a Research Tissue Bank by the National Research Ethics Service. All participants have given informed consent. Details on recruitment and cohort setup have been thoroughly described in a previous publication.^57^ Post-mortem brains underwent full neuropathological dissection, sampling, and characterization by experienced neuropathologists in each of the five network brain banks using a standardized BDR protocol which was based on the BrainNet Europe initiative.^52^ Additional information on the BDR DNA methylation dataset can be found in Shireby et al.^30^

### Genotyping

All NBB samples were genotyped using the NeuroChip Consortium Array (Illumina, San Diego, CA USA).^29^ Quality control was carried out in PLINK 1.9.^58^ Quality control included filtering of variants and individuals based on call rate (< 0.95), Hardy-Weinberg equilibrium (p < 0.000001), relatedness (pi-hat > 0.125) and excess heterozygosity (> 4SD from mean) as well as sex-check and ancestry assessment based on principal component plots. Samples failing genotyping QC were excluded from all analyses. Genotypes were imputed using the Michigan Imputation Server^59^ using default settings and reference data from the Haplotype Reference Consortium^60^, and SNPs with an imputation r^2^ < 0.3 were filtered out. The NeuroChip array has also been designed to allow for screening for known pathogenic mutations in relevant Mendelian neurodegenerative genes. We identified no carriers of definitely or probably pathogenic variants in *SNCA, LRRK2*, and *VPS35* in our discovery sample set.

### DNA methylation analyses, data normalization and quality control

500ng DNA from each sample was bisulfite treated and assessed using the Illumina Infinium MethylationEPIC BeadChip, assigning samples randomly to arrays, including technical replicates for 30 samples. All data processing was performed using R 4.0.3. Raw signal intensity data were imported into R using the minfi^61^ package. For rigorous quality control, we applied a series of checks and filtering steps taking advantage of the minfi and wateRmelon^62^ R packages. CpG sites with a beadcount < 3 in 5% of samples or detection p-value > 0.05 in 1% of samples were filtered out using the *pfilter* function in the wateRmelon package. No samples had detection p-value > 5% in 1% of sites. Two samples were removed due to low median signal intensities as flagged by the minfi *getQC* function. No outliers were detected by the wateRmelon *outlyx* function. Sex chromosome CpGs were used to estimate sample sex and two samples failing sex-check were removed. We estimated the proportion of NeuN positive cells in each sample using reference data from flow sorted frontal cortex cell populations^63^ as implemented in the minfi package.^64^ Two outliers samples with low NeuN proportions were removed from the dataset. We evaluated different normalization methods, ultimately selecting the wateRmelon *dasen* method^62^, which generated the minimum mean difference in beta value across technical replicate pairs in our data and has been successfully employed in previous AD EWAS with similar study design. The normalized *MethylSet* data object was mapped to the genome and probes on sex-chromosomes, probe sequences containing SNPs of any minor allele frequency in the MethylationEPIC annotation and previously reported cross-reactive probes^65^ were filtered out. It has been shown that a considerable proportion of methylation array CpGs have large measurement errors, making them unsuitable for statistical association testing in complex disorders. Taking advantage of the technical replicates we used the CpGFilter package to compute the intra-class correlation coefficient (ICC), which characterizes the relative contribution of the biological variability to the total variability for each probe.^66^ The authors of CpGFilter recommend discarding all probes with ICC below the median. We chose, however, to only filter out the lowest quartile. Finally, the *MethylSet* data object was converted to beta values and the wateRmelon *pwod* function was used to filter out values lying more than four times the interquartile range from the mean, outliers assumed to result from rare SNP artifacts.

### Statistical analyses

There is no general consensus on whether to use beta or M values for statistical analyses of methylation array data. We used beta values for our main analyses because this method has been favored in previously published studies on AD neuropathology that inspired our work. However, we also repeated the statistical analyses with M values, confirming that the main results obtained were highly similar. We generated principal components (PC) and found that after the 5^th^ PC, each PC explained less than 1% of the variation. We observed that NeuN positive cell type proportion and bisulfite conversion experiment plate were associated with PCs, and these were included as covariates in the final analyses in addition to sex, age at death and postmortem delay as recorded by the brain bank. To further minimize the risk of unknown batch effects affecting the results, we estimated surrogate variables (SVs) using the sva package.^67^ In line with previous work, we evaluated models with different numbers of SVs and observed the effects on the test statistic distribution.^10^ We found that including 3 SVs led the inflation measure lambda to fall below 1.2, without further improvement when additional SVs were added. Linear regression was performed using the limma package.^68^ The final lambda value for the discovery analysis was 1.177, yet fell to 1.077 using the *bacon* method developed specifically for epigenome- and transcriptome-wide studies.^69^ This is well in line with previous studies on AD neuropathology^14^, as experience indicates that the low lambda values typically seen in high-quality GWAS cannot be expected in methylation studies. In the main analysis we adjusted for multiple testing using the Benjamini-Hochberg false discovery rate method^70^ which is recommended as default for microarray studies in the limma package. Despite being somewhat less conservative than the Bonferroni method, we considered this to be justified given our two-stage design with independent replication. Fixed-effect meta-analysis was performed using the meta package.^71^ For visualization, we calculated the residuals from a linear regression model with covariates only, excluding Braak stage. The same residuals were used as phenotype in linear regression mQTL analysis in PLINK 1.9. We applied Bonferroni-correction locus by locus to correct for multiple testing in the mQTL assessment.

For the replication cohort, linear regression models against Braak Lewy body stages were run controlling for age, sex, experiment plate and derived cellular proportions. Cell proportions were derived using an algorithm which classifies the cellular populations in the cortex into three proportions (neuronal enriched, oligodendrocyte enriched and microglial enriched) (Shireby et al., manuscript in preparation). Two of the three proportions (neuronal enriched and microglial enriched) were included in the model to eliminate the effects of multi-collinearity.

## Data Availability

Following peer-reviewed publication, genotype and methylation data for the Netherlands Brain Bank cohort will be made publicly available through the Netherlands Neurogenetics Database (https://www.brainbank.nl/nnd-project/). Data from the Brains for Dementia Research cohort are available through application to the Medical Research Council UK Brain Banks Network (https://mrc.ukri.org/brain-banks/) and Dementias Platform UK (https://portal.dementiasplatform.uk).

https://mrc.ukri.org/brain-banks/

https://portal.dementiasplatform.uk

## Acknowledgements

The study was funded by the Norwegian Health Association (grant 4799) and the Research Council of Norway. L.P. and M.T: also received additional funding from the South-Eastern Regional Health Authority, Norway, the Norwegian Parkinson Research Fund and Reberg’s Legacy. WB received funding from the Dutch Parkinson association, Health Holland and Rotary Club Aalsmeer-Uithoorn. The Brains for Dementia Research cohort, including the generation of DNA methylation data, is jointly funded by the UK Alzheimer’s Society and Alzheimer’s Research UK.

## Author contributions

LP, WB and MT designed the study. HG, AJMR and WB provided clinical and neuropathological data for the NBB cohort. SPH and LP performed wetlab work. LP performed analyses and drafted the manuscript. GS performed cohort-level analyses on the BDR data. JAT contributed to data handling and visualization. GS, EH, PF, AJT, SL and JM organized and/or performed collection and generation of BDR data. JM, WB and MT provided supervision. All authors contributed to critical revision of the manuscript.

---

**Supplementary Table 1.** Sample demographics.

| Dataset | Neuropathological diagnosis | N | Mean age at death | Min age | Max age | Prop. Female | Mean PMD min | Min PMD min | Max PMD min |
| --- | --- | --- | --- | --- | --- | --- | --- | --- | --- |
| Discovery sample<br>NBB | Control | 73 | 80.3 | 49 | 99 | 0.70 | 451 | 215 | 1335 |
|  | iLBD | 29 | 85.9 | 72 | 98 | 0.52 | 400 | 150 | 705 |
|  | PD | 139 | 77.7 | 56 | 96 | 0.35 | 360 | 170 | 770 |
|  | DLB | 81 | 78.6 | 54 | 98 | 0.43 | 303 | 190 | 475 |
| Replication sample<br>BDR | Control | 148 | 83.3 | 41 | 104 | 0.50 | 3310 | 390 | 9300 |
|  | iLBD | 18 | 85.5 | 70 | 102 | 0.28 | 2837 | 540 | 7500 |
|  | LBD | 53 | 83.5 | 61 | 97 | 0.40 | 3658 | 480 | 9540 |

PMD = Post mortem delay, NBB = Netherlands Brain Bank, BDR = UK Brains for Dementia Research, DLB = Dementia with Lewy Bodies, iLBD = incidental Lewy Body Disease, PD = Parkinson's disease, LBD = Lewy body dementias

**Supplementary Figure 1.**
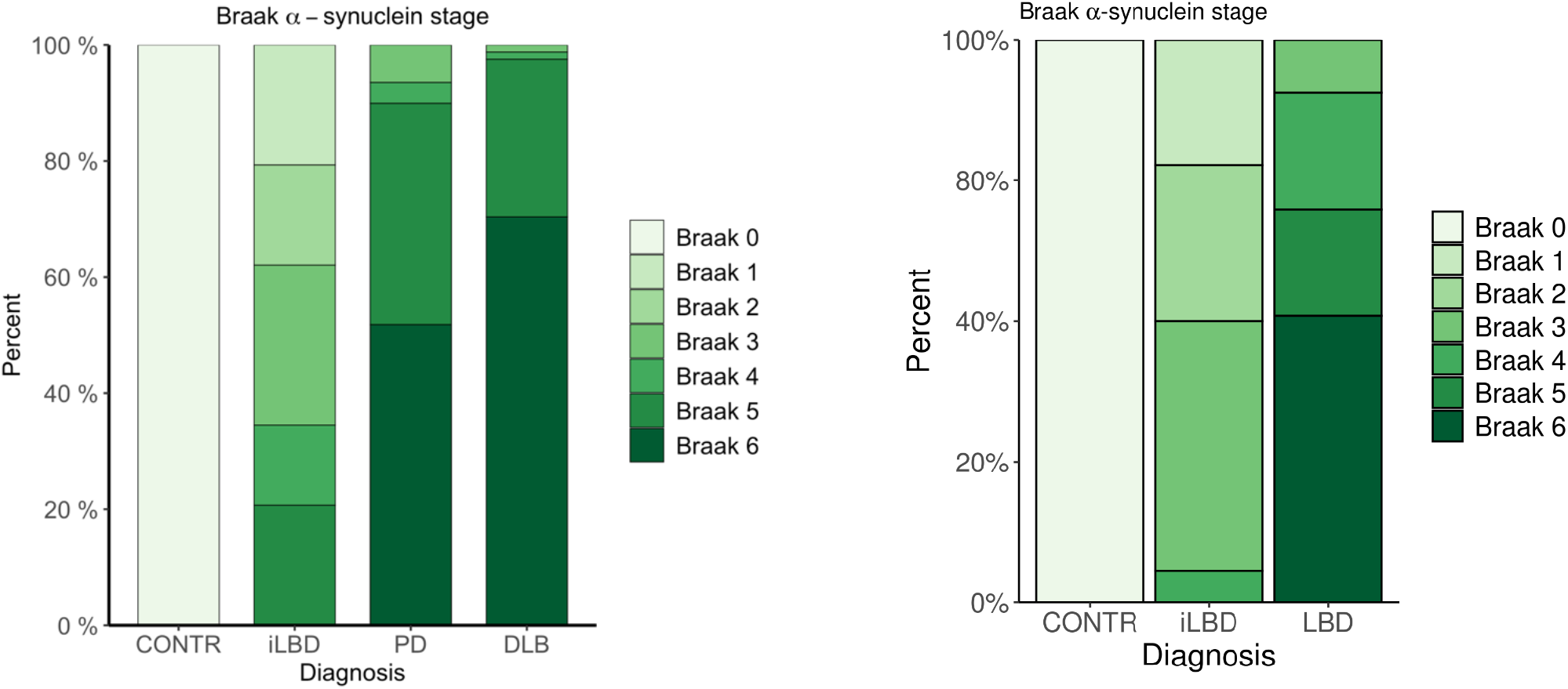
Distribution of Braak Lewy body stages across neuropathological diagnoses. Distribution of Braak α-synuclein stages is shown for the discovery (left) and replication sample (right).

**Supplementary Table 2.** Probes passing FDR threshold in the discovery phase.

| Probe | chr | b37 position | EPIC manifest gene annotation | Beta NBB % | SE NBB % | P NBB | FDR NBB | Beta BDR % | SE BDR % | P BDR | Beta meta % | SE meta % | P meta |
| --- | --- | --- | --- | --- | --- | --- | --- | --- | --- | --- | --- | --- | --- |
| cg01920334 | 21 | 48087978 |  | -0,37 | 0,062 | 5,1E-09 | 0,0030 | -0,04 | 0,111 | 0,716 | -0,29 | 0,054 | 6,1E-08 |
| cg06653480 | 22 | 45034765 |  | 0,58 | 0,100 | 1,1E-08 | 0,0031 | 0,05 | 0,184 | 0,771 | 0,46 | 0,088 | 1,3E-07 |
| cg19772267 | 14 | 63754790 | RHOJ | -0,36 | 0,063 | 2,3E-08 | 0,0044 | 0,04 | 0,123 | 0,770 | -0,28 | 0,056 | 7,6E-07 |
| cg07837822 | 13 | 109740415 | MYO16 | -0,75 | 0,137 | 6,8E-08 | 0,0100 | -0,26 | 0,180 | 0,156 | -0,57 | 0,109 | 1,6E-07 |
| cg07107199 | 1 | 205215911 | TMCC2 | 0,40 | 0,074 | 1,2E-07 | 0,0144 | 0,23 | 0,137 | 0,093 | 0,36 | 0,065 | 2,9E-08 |
| cg14511218 | 10 | 6962843 |  | 0,54 | 0,103 | 2,9E-07 | 0,0252 | 0,35 | 0,159 | 0,030 | 0,48 | 0,087 | 2,5E-08 |
| cg02673002 | 11 | 74459317 | RNF169 | -0,48 | 0,092 | 3,0E-07 | 0,0252 | -0,10 | 0,115 | 0,390 | -0,33 | 0,072 | 3,8E-06 |
| cg04147843 | 11 | 32085613 |  | -0,42 | 0,080 | 3,8E-07 | 0,0258 | -0,11 | 0,216 | 0,605 | -0,38 | 0,075 | 5,0E-07 |
| cg15586054 | 6 | 166072030 | PDE10A | 0,27 | 0,053 | 4,3E-07 | 0,0258 | 0,13 | 0,090 | 0,149 | 0,24 | 0,046 | 2,4E-07 |
| cg16821230 | 3 | 53647443 | CACNA1D | -0,43 | 0,085 | 4,7E-07 | 0,0258 | -0,17 | 0,159 | 0,293 | -0,37 | 0,075 | 5,2E-07 |
| cg09985192 | 14 | 32797255 | AKAP6 | 0,48 | 0,094 | 5,4E-07 | 0,0258 | 0,31 | 0,188 | 0,097 | 0,44 | 0,084 | 1,1E-07 |
| cg11700456 | 12 | 1849881 | ADIPOR2 | 0,42 | 0,083 | 6,3E-07 | 0,0258 | 0,25 | 0,128 | 0,054 | 0,37 | 0,070 | 1,1E-07 |
| cg19793404 | 3 | 87842899 |  | -0,40 | 0,080 | 6,7E-07 | 0,0258 | -0,07 | 0,136 | 0,622 | -0,32 | 0,069 | 3,9E-06 |
| cg22516725 | 16 | 79071140 | WWOX | -0,41 | 0,081 | 6,7E-07 | 0,0258 | -0,22 | 0,135 | 0,099 | -0,36 | 0,069 | 2,1E-07 |
| cg11883790 | 3 | 194877106 | C3orf21 | 0,45 | 0,089 | 6,8E-07 | 0,0258 | -0,04 | 0,148 | 0,774 | 0,32 | 0,077 | 3,0E-05 |
| cg23793336 | 4 | 172509909 |  | 0,74 | 0,147 | 7,1E-07 | 0,0258 | -0,18 | 0,215 | 0,414 | 0,45 | 0,122 | 2,2E-04 |
| cg16701863 | 9 | 116884213 |  | 0,44 | 0,087 | 8,1E-07 | 0,0278 | -0,15 | 0,142 | 0,304 | 0,28 | 0,074 | 1,9E-04 |
| cg04011470 | 8 | 22079330 | PHYHIP | -0,45 | 0,091 | 1,2E-06 | 0,0387 | -0,27 | 0,132 | 0,042 | -0,39 | 0,075 | 1,8E-07 |
| cg19898425 | 14 | 105782464 | BRF1;PACS2 | 0,52 | 0,106 | 1,4E-06 | 0,0425 | 0,32 | 0,164 | 0,049 | 0,46 | 0,089 | 2,1E-07 |
| cg13115910 | 1 | 55840880 |  | -0,37 | 0,076 | 1,5E-06 | 0,0425 | -0,07 | 0,132 | 0,611 | -0,30 | 0,066 | 6,8E-06 |
| cg13560398 | 4 | 129703692 |  | 0,39 | 0,079 | 1,6E-06 | 0,0434 | 0,12 | 0,142 | 0,412 | 0,32 | 0,069 | 3,2E-06 |
| cg25790120 | 12 | 53086242 | KRT77 | 0,44 | 0,092 | 1,9E-06 | 0,0472 | -0,01 | 0,139 | 0,944 | 0,31 | 0,077 | 6,3E-05 |
| cg05516842 | 2 | 25551630 | DNMT3A;MIR1301 | 0,35 | 0,072 | 1,9E-06 | 0,0472 | 0,22 | 0,135 | 0,108 | 0,32 | 0,064 | 4,9E-07 |
| cg13986157 | 3 | 186060255 | DGKG | -0,35 | 0,072 | 2,0E-06 | 0,0480 | -0,31 | 0,135 | 0,022 | -0,34 | 0,063 | 8,7E-08 |

NBB = Netherlands Brain Bank, BDR = UK Brains for Dementia Research, SE = standard error, FDR = false discovery rate by the Benjamini-Hochberg method. Association statistics are based on linear regression adjusting for covariates as described in the main text. Reported meta-analysis results correspond to the inverse-variance fixed effects method. The table is sorted by lowest p-value in the discovery dataset.

**Supplementary Table 3.** Probes passing FDR threshold in meta-analysis with consistent direction of effect and p < 0.05 in both datasets.

| Probe | chr | b37 position | EPIC manifest gene annotation | Beta NBB % | SE NBB % | P NBB | Beta BDR % | SE BDR % | P BDR | Beta meta % | SE meta % | P meta | FDR meta |
| --- | --- | --- | --- | --- | --- | --- | --- | --- | --- | --- | --- | --- | --- |
| cg14511218 | 10 | 6962843 |  | 0,54 | 0,10 | 2,9E-07 | 0,35 | 0,16 | 0,0301 | 0,48 | 0,09 | 2,5E-08 | 0,0083 |
| cg03318382 | 22 | 19691974 |  | 0,46 | 0,10 | 1,8E-05 | 0,52 | 0,16 | 0,0011 | 0,48 | 0,09 | 5,0E-08 | 0,0086 |
| cg13986157 | 3 | 186060255 | DGKG | -0,35 | 0,07 | 2,0E-06 | -0,31 | 0,13 | 0,0216 | -0,34 | 0,06 | 8,7E-08 | 0,0089 |
| cg04011470 | 8 | 22079330 | PHYHIP | -0,45 | 0,09 | 1,2E-06 | -0,27 | 0,13 | 0,0422 | -0,39 | 0,08 | 1,8E-07 | 0,0094 |
| cg19898425 | 14 | 105782464 | BRF1;PACS2 | 0,52 | 0,11 | 1,4E-06 | 0,32 | 0,16 | 0,0487 | 0,46 | 0,09 | 2,1E-07 | 0,0094 |
| cg16231739 | 4 | 122107273 | TNIP3 | -0,28 | 0,06 | 1,6E-05 | -0,31 | 0,11 | 0,0063 | -0,29 | 0,06 | 2,5E-07 | 0,0094 |
| cg11761535 | 3 | 149051031 | TM4SF18 | 0,41 | 0,09 | 4,8E-06 | 0,34 | 0,15 | 0,0230 | 0,39 | 0,08 | 2,5E-07 | 0,0094 |
| cg04381957 | 3 | 16550461 | RFTN1 | -0,40 | 0,08 | 3,9E-06 | -0,26 | 0,13 | 0,0444 | -0,35 | 0,07 | 5,2E-07 | 0,0148 |
| cg21081878 | 21 | 38334730 | HLCS | -0,33 | 0,08 | 2,8E-05 | -0,32 | 0,13 | 0,0114 | -0,33 | 0,07 | 7,6E-07 | 0,0183 |
| cg10961323 | 1 | 24127976 | GALE | 0,44 | 0,10 | 3,7E-05 | 0,43 | 0,16 | 0,0086 | 0,43 | 0,09 | 7,7E-07 | 0,0183 |
| cg25627956 | 12 | 1849936 | ADIPOR2 | 0,31 | 0,08 | 5,4E-05 | 0,37 | 0,13 | 0,0053 | 0,32 | 0,07 | 7,8E-07 | 0,0183 |
| cg07985890 | 11 | 65190999 | NEAT1 | 0,46 | 0,11 | 1,5E-05 | 0,44 | 0,20 | 0,0258 | 0,46 | 0,09 | 8,8E-07 | 0,0195 |
| cg04582365 | 10 | 59155846 |  | 0,46 | 0,11 | 1,8E-05 | 0,39 | 0,17 | 0,0201 | 0,44 | 0,09 | 9,0E-07 | 0,0195 |
| cg08707135 | 17 | 33413689 | RFFL;RAD51L3-RFFL | 0,33 | 0,08 | 1,5E-05 | 0,32 | 0,14 | 0,0281 | 0,33 | 0,07 | 9,3E-07 | 0,0195 |
| cg02059867 | 17 | 38334623 | RAPGEFL1 | -0,24 | 0,06 | 7,4E-05 | -0,32 | 0,11 | 0,0049 | -0,26 | 0,05 | 1,1E-06 | 0,0219 |
| cg26333159 | 11 | 119471437 |  | 0,24 | 0,06 | 2,4E-04 | 0,37 | 0,11 | 0,0014 | 0,27 | 0,06 | 1,4E-06 | 0,0248 |
| cg08206657 | 10 | 14575436 | FAM107B | 0,47 | 0,12 | 8,8E-05 | 0,39 | 0,15 | 0,0088 | 0,44 | 0,09 | 2,1E-06 | 0,0360 |
| cg17930023 | 14 | 94209842 | PRIMA1 | 0,29 | 0,07 | 1,2E-04 | 0,36 | 0,13 | 0,0061 | 0,31 | 0,07 | 2,1E-06 | 0,0360 |
| cg12663253 | 5 | 67522532 | PIK3R1 | -0,26 | 0,06 | 6,7E-05 | -0,32 | 0,13 | 0,0128 | -0,27 | 0,06 | 2,2E-06 | 0,0366 |
| cg01849164 | 10 | 72688074 |  | 0,25 | 0,06 | 3,1E-05 | 0,19 | 0,09 | 0,0280 | 0,23 | 0,05 | 2,3E-06 | 0,0366 |
| cg04511424 | 2 | 113587022 |  | -0,34 | 0,08 | 4,4E-05 | -0,36 | 0,16 | 0,0255 | -0,35 | 0,07 | 2,6E-06 | 0,0393 |
| cg23752007 | 7 | 37429500 | ELMO1 | 0,25 | 0,08 | 1,8E-03 | 0,54 | 0,14 | 0,0001 | 0,33 | 0,07 | 2,7E-06 | 0,0405 |
| cg23076547 | 7 | 34078932 | BMPER | -0,35 | 0,09 | 6,2E-05 | -0,32 | 0,14 | 0,0214 | -0,34 | 0,07 | 3,2E-06 | 0,0436 |
| cg27391403 | 13 | 113416621 | ATP11A | 0,32 | 0,08 | 1,5E-04 | 0,38 | 0,14 | 0,0082 | 0,33 | 0,07 | 3,3E-06 | 0,0446 |
| cg02936493 | 7 | 73223935 |  | 0,22 | 0,05 | 5,7E-05 | 0,29 | 0,13 | 0,0229 | 0,23 | 0,05 | 3,5E-06 | 0,0453 |
| cg22608646 | 17 | 76565037 | DNAH17 | 0,21 | 0,06 | 3,2E-04 | 0,20 | 0,07 | 0,0045 | 0,21 | 0,04 | 3,8E-06 | 0,0453 |
| cg03587422 | 19 | 36367330 | APLP1 | 0,24 | 0,06 | 1,6E-04 | 0,24 | 0,09 | 0,0111 | 0,24 | 0,05 | 4,4E-06 | 0,0471 |
| cg15321814 | 17 | 27387766 |  | -0,17 | 0,04 | 4,8E-05 | -0,14 | 0,07 | 0,0396 | -0,16 | 0,04 | 4,5E-06 | 0,0471 |

NBB = Netherlands Brain Bank, BDR = UK Brains for Dementia Research, SE = standard error, FDR = false discovery rate by the Benjamini-Hochberg method. Association statistics are based on linear regression adjusting for covariates as described in the main text. Reported meta-analysis results correspond to the inverse-variance fixed effects method. The table is sorted by lowest p-value in the meta-analysis.

